# Evaluating the Performance of ChatGPT-4o Vision Capabilities on Image-Based USMLE Step 1, Step 2, and Step 3 Examination Questions

**DOI:** 10.1101/2024.06.18.24309092

**Authors:** Avi A. Gajjar, Harshitha Valluri, Tarun Prabhala, Amanda Custozzo, Alan S Boulos, John C. Dalfino, Nicholas C. Field, Alexandra R. Paul

## Abstract

**Introduction:** Artificial intelligence (AI) has significant potential in medicine, especially in diagnostics and education. ChatGPT has achieved levels comparable to medical students on text-based USMLE questions, yet there’s a gap in its evaluation on image-based questions.

**Methods:** This study evaluated ChatGPT-4’s performance on image-based questions from USMLE Step 1, Step 2, and Step 3. A total of 376 questions, including 54 image-based, were tested using an image-captioning system to generate descriptions for the images.

**Results:** The overall performance of ChatGPT-4 on USMLE Steps 1, 2, and 3 was evaluated using 376 questions, including 54 with images. The accuracy was 85.7% for Step 1, 92.5% for Step 2, and 86.9% for Step 3. For image-based questions, the accuracy was 70.8% for Step 1, 92.9% for Step 2, and 62.5% for Step 3. In contrast, text-based questions showed higher accuracy: 89.5% for Step 1, 92.5% for Step 2, and 90.1% for Step 3. Performance dropped significantly for difficult image-based questions in Steps 1 and 3 (p=0.0196 and p=0.0020 respectively), but not in Step 2 (p=0.9574). Despite these challenges, the AI’s accuracy on image-based questions exceeded the passing rate for all three exams.

**Conclusions:** ChatGPT-4 can handle image-based USMLE questions above the passing rate, showing promise for its use in medical education and diagnostics. Further development is needed to improve its direct image processing capabilities and overall performance.

## INTRODUCTION

Artificial intelligence (AI) has emerged as a transformative technology in medicine, offering significant potential to enhance clinical practice, diagnostics, and medical education.^1,2^ The deployment of AI models in various domains of healthcare has demonstrated promising results, particularly in the realm of natural language processing (NLP) for medical question answering. Among these, large language models such as Chat Generative Pre-trained Transformer (ChatGPT) have shown considerable success by achieving performance levels comparable to medical students on standardized exams like the United States Medical Licensing Examination (USMLE).^3–5^

Numerous studies have explored the capabilities of AI in handling medical board examinations. These investigations have primarily focused on text-based questions, consistently demonstrating that AI can provide accurate and contextually relevant answers.^6–8^ For example, recent research indicates that ChatGPT surpasses previous models in logical justification and coherence when responding to these exams’ questions.^1,9^ However, a critical limitation of these studies is the exclusion of image-based questions, which form an integral part of medical education and clinical assessment.

Medical licensing exams frequently include visual data such as radiographs, histopathology slides, and clinical photographs to test competency in interpreting diagnostic images.^6^ To date, no studies have comprehensively evaluated AI models’ performance on these image-based questions, leaving a significant gap in understanding the full capabilities and limitations of AI in the medical field.

This study aims to address this gap by investigating whether AI can handle images in addition to text, thereby providing a more comprehensive evaluation of its potential in medical applications. Specifically, we will assess the performance of AI models on image-based questions from a standardized medical question bank.

## METHODS

### Study Design

This study evaluates the performance of ChatGPT-4, specifically its ChatGPT-4o version, on USMLE image-based questions. The objective is to determine whether the AI can effectively interpret and respond to image-embedded queries, thereby exploring the potential for multimodal AI applications in medical education.

We utilized a comprehensive set of test questions designed to evaluate AI performance on USMLE Step 1 and Step 2 exams. These questions were sourced from a joint program of the Federation of State Medical Boards of the United States, Inc. (FSMB), and the National Board of Medical Examiners® (NBME®).^10–12^

### Data Preparation

To prepare each image-based question for input into ChatGPT, we employed an image-captioning system due to ChatGPT’s current limitations in directly processing visual data. This involved generating captions for the images using convolutional neural networks (CNNs) trained to produce medical image descriptions. These captions were then integrated into the question prompts, accompanied by the original question text and multiple-choice options.

### Prompt Engineering

Prompt engineering was standardized to ensure consistency in AI responses. Each question was formatted to include an image description, followed by the original question text and multiple-choice options. For example, the prompt might read:

“The following image shows a chest radiograph with bilateral patchy infiltrates. Based on the information provided in the image, answer the following question: What is the most likely diagnosis? A) Tuberculosis B) Pneumonia C) Lung cancer D) Pulmonary embolism E) Sarcoidosis.”

The testing of the ChatGPT-4o version involved manually entering each question into the ChatGPT interface as a new prompt, ensuring no memory carryover between successive questions. The model’s answer to each question was documented and analyzed.

### Statistical Analysis

All statistical analyses were conducted using Stata software (version 18.0 SE; StataCorp LLC, College Station, TX, USA). Independent t-tests were applied to determine the significance of performance differences across question difficulty levels and between correct and incorrect responses. A threshold of P<.05 was set for determining statistical significance. The study did not involve human subjects or patient data and therefore did not require ethical approval. All test questions were sourced from publicly available or licensed question banks.

## RESULTS

### Overall Performance

The overall performance of ChatGPT-4 on USMLE Step 1, Step 2, and Step 3 was evaluated. The data set included 376 questions in total, comprising 54 questions with images. Specifically, 11.6% of USMLE Step 2 questions featured images, and up to 20.2% of USMLE Step 1 questions included visual data.

The accuracy for Step 1 was 85.71%, Step 2 was 92.50%, and Step 3 was 86.86%. When comparing text-based and image-based questions, significant differences were observed. For Step 1, the accuracy for image-based questions was 70.83%, while text-based questions achieved 89.47%. In Step 2, the accuracy was consistent for both types of questions, with 92.86% for image-based and 92.45% for text-based questions. However, in Step 3, image-based questions had an accuracy of 62.50%, compared to 90.08% for text-based questions.

### Performance by Question Type

For text-based questions, ChatGPT-4 showed high accuracy across all steps: 89.47% for Step 1, 92.45% for Step 2, and 90.08% for Step 3. Conversely, image-based questions presented a greater challenge. Step 1 achieved an accuracy of 70.83%, Step 2 reached 92.86%, and Step 3 managed 62.50%. These results highlight a noticeable performance gap in Steps 1 and 3 compared to text-based questions.

The analysis of performance by difficulty level for image-based questions revealed significant drops in accuracy for more difficult questions. T-tests indicated notable differences in performance between easy and difficult image-based questions in Steps 1 and 3. For Step 1, there was a significant performance disparity (p=0.0196), and Step 3 also exhibited a significant difference (p=0.0020). In contrast, Step 2 maintained high accuracy across all difficulty levels, showing no significant difference (p=0.9574).

## DISCUSSION

This study provides important insights into the performance of AI, particularly ChatGPT-4, on the USMLE image-based questions. Previous literature has extensively evaluated AI’s capabilities in answering text-based medical questions, demonstrating proficiency levels comparable to human medical students.^3,13^ However, there has been a considerable gap in assessing AI’s competencies on image-based questions, which are critical for diagnosing and understanding medical conditions. By addressing this gap, our study offers a comprehensive evaluation and highlights the potential utility and limitations of AI in medical education and diagnostic applications.

### Performance Disparity between Text-Based and Image-Based Questions

Our results revealed a notable performance disparity between text-based and image-based questions on the USMLE. ChatGPT-4 demonstrated high accuracy for text-based questions, attaining 89.47% for Step 1, 92.45% for Step 2, and 90.08% for Step 3. These findings are consistent with prior studies where AI models have shown significant capability in handling text-based inquiries, often surpassing human benchmarks.^3,13^ However, the accuracy for image-based questions was considerably lower, with 70.83% for Step 1, 92.86% for Step 2, and 62.50% for Step 3. The largest discrepancies were observed in Steps 1 and 3, underscoring the difficulties AI faces in interpreting complex visual data. These results underscore a critical need to enhance AI’s image-processing systems to achieve more balanced performance across different question types.

### Challenges in Interpreting Visual Data

Our analysis highlighted specific challenges AI faces in interpreting visual data. Despite not categorizing errors into types, common issues included AI’s difficulty in recognizing fine details and accurately interpreting complex medical images. These difficulties often led to incorrect or incomplete diagnoses, reflecting a significant gap in AI capabilities compared to human experts. The problems were most pronounced in Step 3 questions, where the accuracy dropped to 62.50%. Advanced image-processing algorithms may be required to improve AI’s proficiency.

Prior studies suggest that incorporating more sophisticated machine learning techniques, like convolutional neural networks (CNNs), might enhance AI’s ability to analyze images more accurately.^14,15^ Moving forward, it will be crucial to develop AI systems that can maintain high accuracy in both text and image-based questions to provide reliable support in medical diagnostics and education.

### Implications for Medical Education and Diagnostics

Enhancing AI capabilities to interpret medical images could revolutionize medical training by providing more accurate feedback on diagnostic questions that involve visual data.^15^ This could improve the learning experience for medical students and ensure they are better prepared for real-world clinical scenarios. Additionally, improved AI performance in diagnostic imaging may augment clinical practice by offering a reliable second opinion, potentially reducing diagnostic errors and improving patient outcomes. The integration of AI into medical education tools has the potential to create a more interactive and engaging learning environment, which could lead to better retention and understanding of complex medical information.^3,16^

### Future Directions

To address the current limitations and enhance AI performance, several recommendations for future research are proposed. There is a need to develop sophisticated algorithms specifically designed for interpreting medical images. Techniques such as convolutional neural networks (CNNs) and generative adversarial networks (GANs) could be leveraged to improve the accuracy and interpretability of AI models.^15^ AI models should be trained on extensive and diverse datasets that include a broad range of medical images and scenarios. This would increase the robustness and generalizability of AI systems across different types of medical imaging modalities and clinical contexts. Integrating text and image data in AI models could provide a more comprehensive approach to medical diagnostics and education. Multimodal AI would leverage the strengths of both data types, resulting in more accurate and contextually relevant outputs, thus enhancing clinical decision-making and learning outcomes.^17^ Collaboration between AI developers and medical educators is essential to ensure the developed AI systems are aligned with practical needs. Such partnerships can refine AI models to make them more applicable and beneficial in real-world medical settings.^16^ By following these recommendations, we can significantly improve the capabilities of AI in medical applications, ensuring it meets the rigorous demands of both educational and clinical environments. Future research should aim to continuously refine and test these AI models to maximize their potential and utility in the medical field.

## CONCLUSIONS

This study demonstrates that ChatGPT-4, particularly in its ChatGPT-4o version, can handle image-based questions at an accuracy that exceeds the passing rate, thereby offering a comprehensive example of its ability to succeed in the USMLE exams. The AI’s performance on both text-based and image-based questions highlights its potential as a tool to assist medical students and professionals. Despite the current limitations in directly processing visual data, ChatGPT-4’s ability to interpret image captions and respond accurately underscores its promise in enhancing medical education and diagnostic processes. These findings suggest that with further development, AI could play a significant role in training and assessment within the medical field, providing robust support for both learners and practitioners.

## Data Availability

All data produced in the present study are available upon reasonable request to the authors

## Abbreviations

AI: Artificial Intelligence
ChatGPT: Chat Generative Pre-trained Transformer
USMLE: United States Medical Licensing Examination
FSMB: Federation of State Medical Boards
NBME: National Board of Medical Examiners
CNN: Convolutional Neural Networks
GAN: Generative Adversarial Networks

## FIGURE LEGENDS

**Figure 1.**
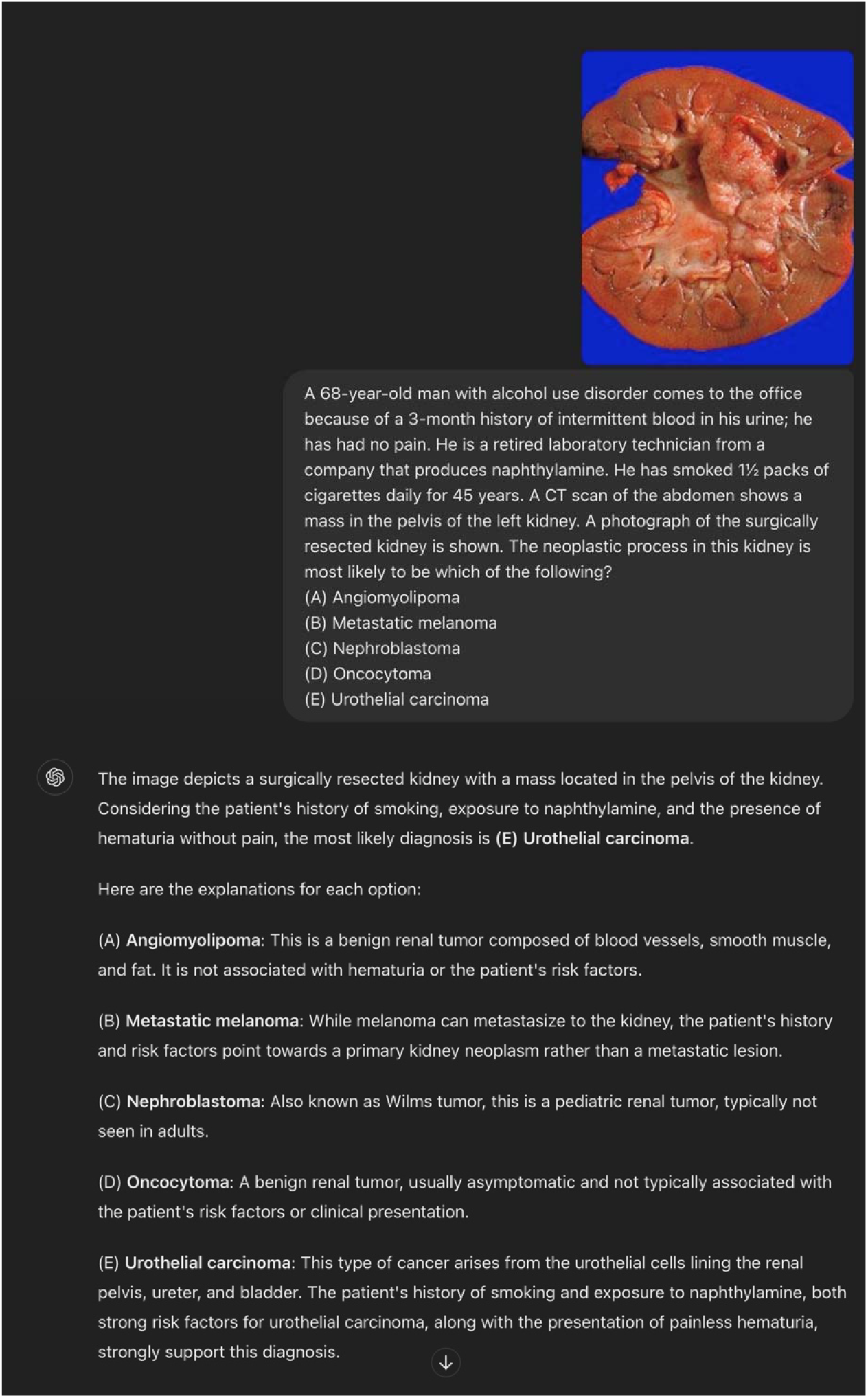
Example of prompted USMLE Step 1 question to ChatGPT

**Figure 2.**
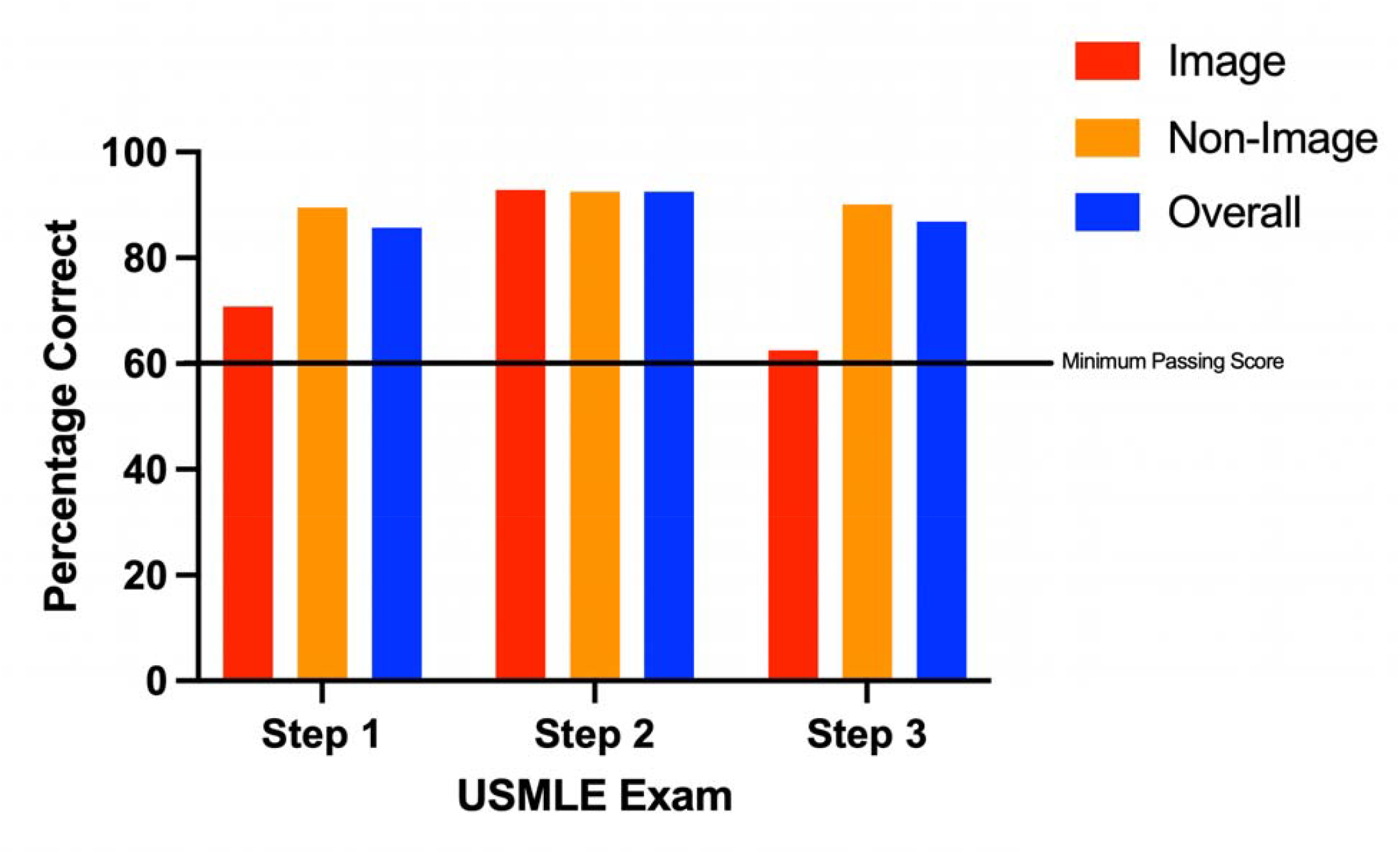
Accuracy of image, non-image, and overall questions for all USMLE Exams.

**Table 1.**
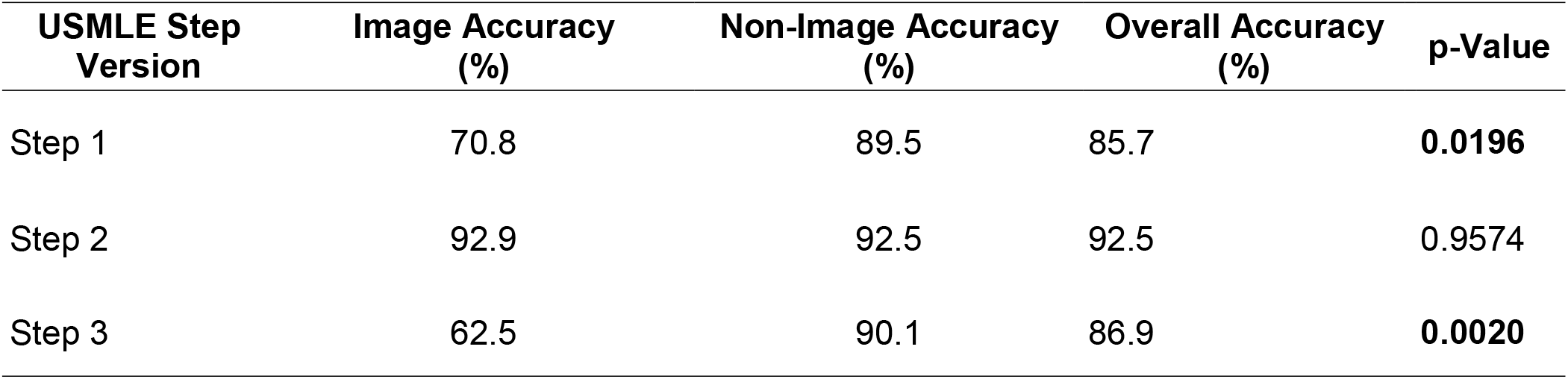
Accuracy Metrics and Statistical Significance for USMLE Steps.

